# Lower help-seeking intentions and subsequent depressive symptoms among adolescents with high autistic traits: A population-based cohort study

**DOI:** 10.1101/2021.02.27.21252566

**Authors:** Mariko Hosozawa, Syudo Yamasaki, Shuntaro Ando, Kaori Endo, Yuko Morimoto, Sho Kanata, Shinya Fujikawa, Noriko Cable, Hiroyasu Iso, Mariko Hiraiwa-Hasegawa, Kiyoto Kasai, Atsushi Nishida

**Affiliations:** Institute for Global Health Policy Research, Bureau of International Health Cooperation, National Center for Global Health and Medicine, Japan; Department of Epidemiology and Public Health, University College London, United Kingdom; Department of Paediatrics and Adolescent Medicine, Juntendo University, Japan; Research Center for Social Science & Medicine, Tokyo Metropolitan Institute of Medical Science, Japan; Department of Neuropsychiatry, Graduate School of Medicine, The University of Tokyo, Japan; Department of Psychology, Ube Frontier University, Japan; Department of Psychiatry, Teikyo University School of Medicine, Japan; Public Health, Department of Social Medicine, Osaka University Graduate School of Medicine, Japan; Department of Evolutionary Studies of Biosystems, The Graduate University for the Advanced Studies, SOKENDAI, Japan

## Abstract

**Background:** Adolescents with high autistic traits in the general population are at increased risk of depression. Despite the importance of help-seeking for early intervention, evidence on help-seeking intentions among this population is scarce.

**Aims:** To examine the help-seeking intentions and preferences for depression by the level of autistic traits in adolescents, and test how help-seeking intentions mediate the association between autistic traits and depressive symptoms.

**Method:** Participants were from the Tokyo Teen Cohort, a population-based cohort in Japan. They were classified into two groups according to parent-rated autistic traits measured by the short-version of the Autism-Spectrum Quotient (≥6 as AQhigh). Help-seeking intentions and preferences were assessed at age 12 using a depression vignette. Depressive symptoms were self-rated at age 14 using the Short Mood and Feelings Questionnaire. Data were analysed using multivariable regression analysis and structural equation modelling.

**Results:** Of the 2,505 adolescents in the study, 200 (8%) were classified as AQhigh. In the AQhigh-group, 40% of the adolescents reported not having help-seeking intentions, although 93% recognised the need and 97% had someone to rely on. Parents of the AQhigh-group also reported fewer help-seeking intentions. The AQhigh-group was associated with an increased risk of not having help-seeking intentions (OR 1.83, 95%CI 1.35–2.49), which explained 19% of the above association.

**Conclusions:** Lower help-seeking intentions among adolescents with high autistic traits partially explained their increased risk for subsequent depressive symptoms. Interventions to promote help-seeking intentions among this population should involve both adolescents and their parents and ideally be provided before adolescence.

## Introduction

### Background

Autism spectrum disorder (hereafter referred to as autism) is a neurodevelopmental condition that is characterised by difficulties in social communication and interaction and restricted, repetitive patterns of behaviour, interests, or activities. (1) Autism is a dimensional condition, and autistic traits exists as a continuum across the general population. (2) Around 70–80% of all people with autism experience mental health problems such as depression, (3) the prevalence of which may increase during adolescence. (4) Furthermore, recent research shows that not only adolescents that are diagnosed with autism but those with high autistic traits in the general population are at increased risk of depression. (5) The evidence highlights the need for approaches that prevent or reduce mental health problems, particularly depression, among those with high autistic traits regardless of whether they have been formally diagnosed.

Help-seeking intentions facilitate early intervention, which is essential for the prevention or reduction of mental health problems. (6) However, qualitative studies have shown that even in countries with good access to health care, autistic individuals report a reluctance towards seeking help for their mental health problems, partly due to their difficulties in recognising the need for help and increased self-reliance. (7, 8) Furthermore, most previous studies on help-seeking and mental health problems were cross-sectional, (9, 10) and there is limited evidence as to whether help-seeking intentions influence subsequent mental health problems, within or outside of the context of autism. Therefore, improving our understanding of help-seeking intentions and their role in preventing subsequent depressive symptoms among individuals with high levels of autistic traits, including those who are undiagnosed, can help develop preventive interventions in this population. Nevertheless, there is a lack of quantitative studies that examine the help-seeking intentions for mental health problems among individuals with high autistic traits, both diagnosed and undiagnosed, in the general population.

### Objectives

Using the Tokyo Teen Cohort (TTC), an ongoing population-based cohort study of adolescents living in Tokyo, our primary aim was to investigate help-seeking intentions related to depression and help-seeking preferences according to the level of autistic traits. We also aimed to examine the potential mediating role of help-seeking intentions in the association between autistic traits and subsequent depressive symptoms in adolescence. We hypothesised that adolescents with high autistic traits would show decreased help-seeking intentions, which would explain their increased risk of subsequent depressive symptoms.

## Methods

### Study population

The TTC is an ongoing population-based cohort study that is following the physiological and psychological development of 3,171 children born between 2002 and 2004 from the age of 10 who are living in three municipalities in the metropolitan area of Tokyo, Japan. The study details are described elsewhere. (11) Of the 3,007 children who took part in the TTC at age 12 (when information on their autistic traits and help-seeking intentions was collected), those without valid information on their help-seeking intentions or autistic traits (n = 482 and 20, respectively) were excluded from our study. The authors assert that all procedures that contributed to this work comply with the ethical standards of the relevant national and institutional committees on human experimentation and with the Helsinki Declaration of 1975, as revised in 2008. All procedures involving human participants were approved by the institutional review boards of the Tokyo Metropolitan Institute of Medical Science (approval number: 12–35), SOKENDAI (Graduate University for Advanced Studies, 2012002), and the University of Tokyo (10057). Written informed consent was obtained from all the parents of the participating children, and informed assent was obtained from the children.

### Involvement of the patients and the public

The participants’ advisory board of the TTC, which consisted of the main carers for the adolescents, provided feedback on the research questions, outcome measures, and design or implementation of the study. None of the board members were asked to advise on the interpretation or writing of the results; however, the results were disseminated to the study participants through their dedicated website: http://ttcp.umin.jp/.

### Participants’ help-seeking intentions for depression at age 12

We measured the participants’ help-seeking intentions for depression at age 12 using a vignette written in Japanese. The vignette presented a boy named ‘Taro’ (a popular boy’s name in Japan) who was meant to display symptoms of depression. The symptoms presented in the vignette covered the Diagnostic and Statistical Manual of Mental Disorders (Fourth Edition) and International Classification Of Diseases 10 criteria for major depression and mimicked those used in other large-scale studies. (12) Hereafter follows an example of the vignette: ‘For the last several weeks, Taro has been feeling unusually sad. He is tired all the time and has trouble sleeping at night. Taro does not feel like eating and has lost weight. He cannot keep his mind on his studies, and his grades have dropped. He puts off making any decisions and even day-to-day tasks, such as studying and extracurricular activities, seem too much for him. His parents and teachers are very concerned about him’. Following the vignette, the children were asked four questions that assessed their help-seeking intentions, which are described below.

#### 1. Recognition of the need for help in the vignette case

Adolescents were asked, ‘What do you think about Taro’s condition?’ The response options were [1] ‘He needs help’; [2] ‘He has a problem, but does not need help’; or [3] ‘He does not have a problem’.

#### 2. Help-seeking intentions for depression

Adolescents were asked to respond to the question, ‘If you were in the same situation as Taro, would you seek help from others?’ The response options were [1] ‘I would consult someone immediately’ or [2] ‘I would wait and see without consulting anyone’, from which we created a binary variable based on their response.

#### 3. The number of people whom the adolescent could rely on for help (I have someone to rely on for help)

Adolescents were asked, ‘If you were in the same situation as Taro, and if you considered asking someone for help, how many people do you think you could rely on for help?’ The participants were given the following options: from 0 to 4 people and 5 or more people. The responses were summarised into a binary variable that represented having no one vs having someone, given that not having anyone to rely on for help had a particular impact on mental health. (13)

#### 4. Assessment of help-seeking preferences

Adolescents were asked, ‘If you were in the same situation as Taro, who would you ask for help?’ Then, they were asked to select each answer that applied in response to the question: ‘Friends’, ‘Family’, ‘Relatives’, ‘Teachers’, ‘School counsellor’, ‘Medical doctor’, or ‘Online forum’. All responses were treated in a binary format (not selected vs selected).

### Autistic traits

The participants’ autistic traits were measured at age 12 (wave 2) using the short form of the Autism-Spectrum Quotient adolescent version, a parent-reported questionnaire that was designed to measure autistic traits in the general population. (14) The questionnaire consisted of 10 items that were given a score of either 0 or 1. We calculated the total score (range 0–10, where higher total scores indicated greater autistic tendencies) and classified the participants that scored above 6 as those with high autistic traits (the AQhigh group), according to the findings from a previous study. (14)

### Depressive symptoms

The participants’ depressive symptoms were measured using the Short Mood and Feelings Questionnaire (SMFQ) that was administered at age 14. (15) The SMFQ is a well-validated 13 item self-reported questionnaire for children and adolescents to evaluate their depressive symptoms in the preceding two weeks (range 0–26; where higher scores indicated more severe depressive conditions). (16) It has been used in several studies of adolescents with autism. (5)

### Covariates

Based on previous studies, the following covariates, all measured at age 10, were included in our model as potential confounders (13): the child’s sex; relative poverty of the household as indicated by an annual household income below 4,000,000 Yen, which is just below the median national income in Japan; mother’s highest level of education (graduated from high school or higher); child’s intelligence quotient (assessed through an interview that used the short-form of the Wechsler Intelligence Scale for Children; details are provided elsewhere) (17); parent-rated emotional symptoms measured using the emotional difficulties sub-scores of the Strengths and Difficulties Questionnaire (range 0–10, where higher scores indicated more severe emotional problems) (18); and parental help-seeking intentions, which were measured using the adult-version of the depression vignette.

### Statistical analyses

We first conducted a descriptive analysis to examine the association between the exposure and study variables. We also conducted a sample bias analysis to examine whether children in our analytic sample differed from those excluded from our study in relation to our study variables. We then applied multivariable logistic and regression analyses to examine the association between 1) autistic traits and help-seeking intentions (logistic) and 2) autistic traits and self-rated depressive symptoms at age 14 (regression). In both analyses, an unadjusted model (Model 1), a model that was adjusted for sex (Model 2), and a model that was further adjusted for potential confounders (i.e., low annual household income, low maternal education, parental help-seeking intentions, child’s intelligence quotient, and parent-rated emotional symptoms [Model 3]). Our preliminary analyses showed no evidence that suggested differences in associations according to sex or level of parent-rated emotional symptoms at age 10; therefore, the combined results are presented. Possible mediation by the child’s help-seeking intentions on the association between autistic traits and depressive symptoms at age 14 was tested in terms of the direct and indirect effects. As part of the sensitivity analysis, we repeated the analyses by excluding adolescents whose parents reported that a diagnosis of autism was made by age 12 (n = 30). Descriptive analyses were conducted using Stata SE version 16.1 for Windows10 (StataCorp, College Station, TX, United States), and all other analyses were conducted using structural equational modelling in Mplus8.5 (Muthén & Muthén, Los Angeles, CA, United States).

### Missing data

The missing data from each variable ranged from 0.04% (from the child’s intelligence quotient) to 27% (from the depressive symptoms at age 14). We imputed missing covariates and outcomes using multiple imputation by chained equations to minimise data loss. (19) Regression analyses were run across 27 imputed datasets and adjusted using Rubin’s rules. The imputed results were broadly similar to those obtained using the observed cases (Tables S1 and S5); therefore, the former are presented here.

## Results

Of the 2,505 adolescents in our study sample, 200 (8.0%) were classified into the AQhigh group (Table 1). Compared to the adolescents in the AQlow group, those in the AQhigh group were more likely to be boys (51.6 vs 68.0%, p < 0.001) or had received a diagnosis of autism (0.7% vs 7.6%, p < 0.001, Table S1). However, the two groups did not differ in terms of the family’s socioeconomic status or children’s mean intelligence quotient. Interestingly, parents of the adolescents in the AQhigh group were less likely to have help-seeking intentions (79.5% vs 71.0%, p = 0.005). At age 14, adolescents in the AQhigh group showed significantly higher mean self-rated depressive symptoms than those in the AQlow group. The results of our sample bias analysis showed that adolescents whose parents reported a diagnosis of autism by age 12 were more likely to be excluded from our analysis (Table S2).

**Table 1.** Descriptive characteristics for the imputed sample (N = 2,505)

|  | AQlow<sup>a</sup><br>(92.0%) | AQhigh<sup>a</sup><br>(8.0%) | *p*-value<sup>b</sup> |
| --- | --- | --- | --- |
| Autistic traits at age 12, mean (SE) | 2.1 (0.0) | 6.7 (0.1) | < 0.001 |
| Sex |  |  |  |
| Male | 51.6 | 68.0 | < 0.001 |
| Female | 48.4 | 32.0 |  |
| Low annual household income<sup>c</sup> |  |  |  |
| No | 89.6 | 88.1 | 0.53 |
| Yes | 10.4 | 11.9 |  |
| Low maternal education<sup>d</sup> |  |  |  |
| No | 84.0 | 81.3 | 0.33 |
| Yes | 16.0 | 18.7 |  |
| Parental help-seeking intentions<sup>e</sup> |  |  |  |
| No | 20.5 | 29.0 | 0.005 |
| Yes | 79.5 | 71.0 |  |
| Child intelligence quotient, mean (SE) | 108.1 (0.3) | 106.8 (1.1) | 0.24 |
| Parent-rated emotional symptoms, mean (SE) | 1.5 (0.0) | 2.5 (0.2) | < 0.001 |
| Self-reported depressive symptoms at age 14, mean (SE) | 3.0 (0.1) | 4.1 (0.4) | 0.007 |

Variables were measured in the first wave of the study (age 10) unless otherwise stated.
<sup>a</sup>Defined as scoring above the suggested clinical-cut off value on the short form of the Autism-Spectrum Quotient adolescent version as measured by a parent at age 12.
<sup>b</sup>*p*-value for group difference obtained from the F statistic tests.
<sup>c</sup>Defined as a household income below 4,000,000 Yen.
<sup>d</sup>Defined as a high school graduate or below.
<sup>e</sup>Measured through the adult version of the depression vignette.
Abbreviations: AQ, Autism-Spectrum Quotient; SE, standard error.

Table 2 shows that a significantly higher proportion of adolescents in the AQhigh group reported not having help-seeking intentions than those in the AQlow group (40.0% vs 26.5%, p < 0.001). However, both the AQhigh and AQlow groups responded similarly regarding the recognition of the need for help in the vignette and having someone to ask for help (92.5% vs 94.4% and 96.5% vs 98.1%, p = 0.50 and p = 0.11, respectively).

**Table 2.** Adolescents’ help-seeking intentions for depression according to the level of autistic traits (N =2,505)

|  | AQlow<br>(n=2,305,<br>92.0%) |  | AQhigh<br>(n=200, 8.0%) |  | *p*-value<sup>a</sup> |
| --- | --- | --- | --- | --- | --- |
|  | n | % | n | % |  |
| Did not have help-seeking intentions | 611 | 26.5 | 80 | 40.0 | < 0.001 |
| Recognition of the need for help in the vignette |  |  |  |  |  |
| He needs help | 2,176 | 94.4 | 185 | 92.5 | 0.50 |
| He has a problem, but does not need help | 100 | 4.3 | 11 | 5.5 |  |
| He does not have a problem | 29 | 1.3 | 4 | 2.0 |  |
| [I have] someone to rely on for help | 2,268 | 98.1 | 192 | 96.5 | 0.11 |

All variables were measured through the depression vignette at age 12.
<sup>a</sup>*p*-value for the group difference obtained from the Chi-square tests.

Fewer adolescents in the AQhigh group reported seeking help from friends compared to the AQlow group (61.9% vs 77.5%, p < 0.001); however, the overall distribution pattern of the help-seeking preferences did not differ between the groups (Figure 1). Adolescents from both groups were most likely to seek help from their family (79.7% and 84.0% in the AQhigh and AQlow groups, respectively), followed by their friends. They were less likely to seek help from professionals such as school counsellors or medical doctors (19.3% vs 24.4% and 10.7% vs 10.8%, respectively).

**Figure 1.**
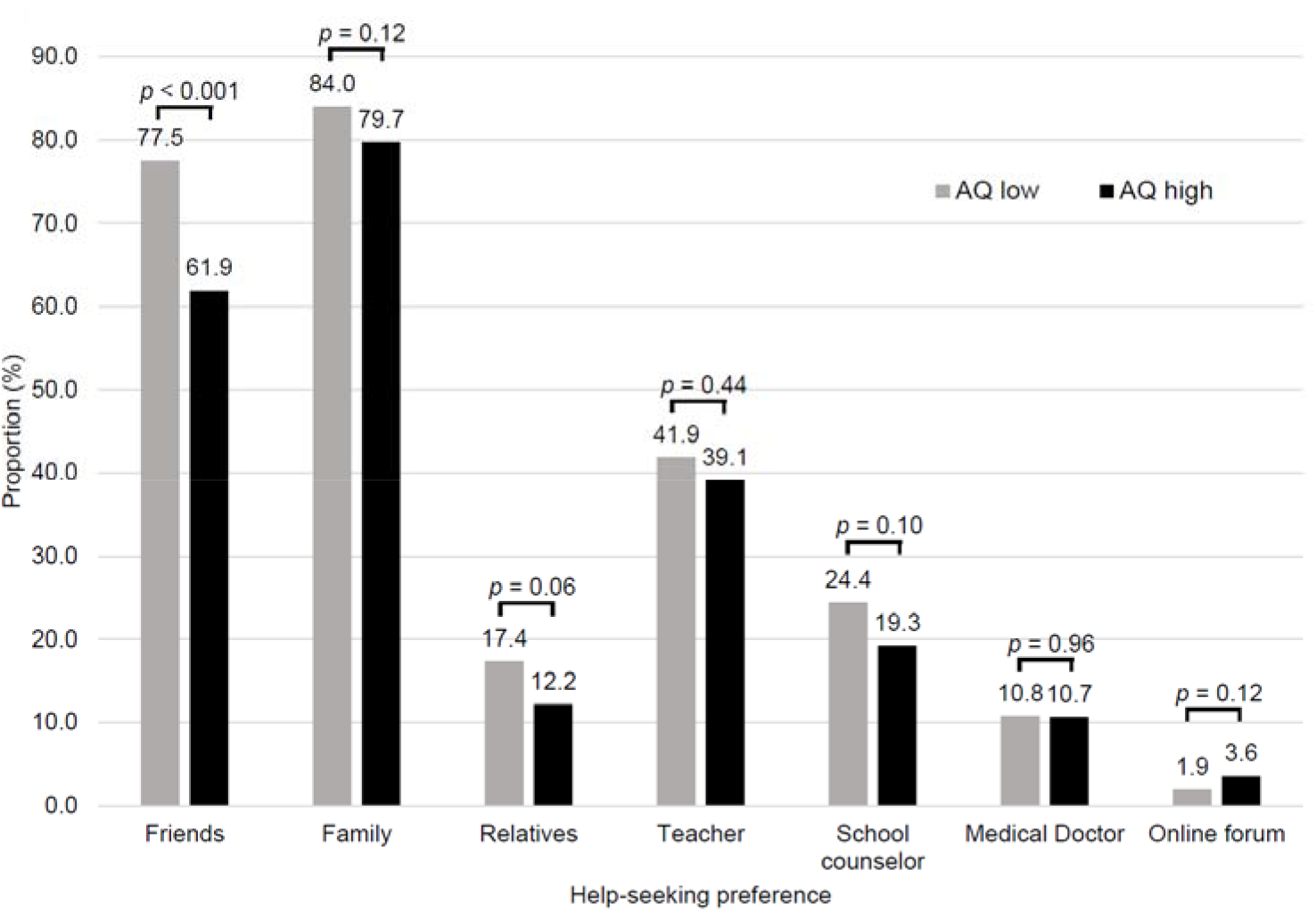
Help-seeking preferences for depression according to the level of autistic traits. The proportion of adolescents that reported seeking help from each source according to the autistic traits, based on observed results. Participants were able to select multiple responses. The *p*-value for the group differences was obtained from Chi-square tests. Abbreviations: AQ, Autism-Spectrum Quotient.

The relationship between autistic traits and not having help-seeking intentions was further confirmed in our multivariable logistic analysis (Table 3). After adjusting for confounders, the odds of the AQhigh group not having help-seeking intentions were 1.84 times greater than the AQlow group (95% CI 1.35–2.50, [Model 3]). Additionally, being in the AQhigh group was associated with significantly higher depressive symptoms (b = 1.04, 0.30–1.78 for Model 3, Table S3). Mediation analysis revealed that not having help-seeking intentions accounted for approximately 19% of the association between AQhigh and depressive symptoms at age 14 (Table S4). Our sensitivity analysis, that excluded children diagnosed with autism at age 12, revealed similar results (Table S6–S7).

**Table 3.**
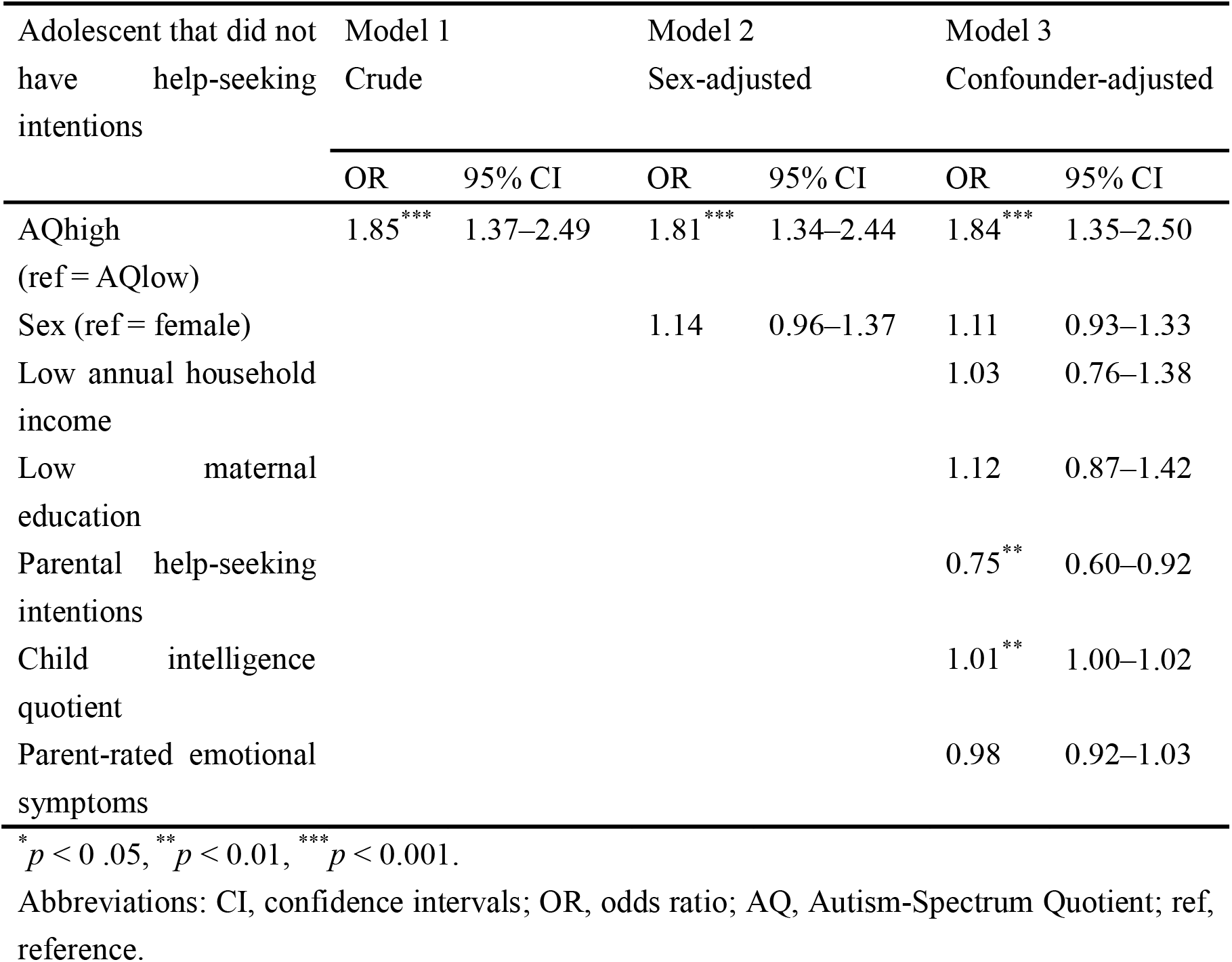
The adolescents’ help seeking intentions according to the level of autistic traits (N = 2,505)

| Adolescent that did not have help-seeking intentions | Model 1<br>Crude |  | Model 2<br>Sex-adjusted |  | Model 3<br>Confounder-adjusted |  |
| --- | --- | --- | --- | --- | --- | --- |
|  | OR | 95% CI | OR | 95% CI | OR | 95% CI |
| AQhigh<br>(ref = AQlow) | 1.85*** | 1.37–2.49 | 1.81*** | 1.34–2.44 | 1.84*** | 1.35–2.50 |
| Sex (ref = female) |  |  | 1.14 | 0.96–1.37 | 1.11 | 0.93–1.33 |
| Low annual household income |  |  |  |  | 1.03 | 0.76–1.38 |
| Low maternal education |  |  |  |  | 1.12 | 0.87–1.42 |
| Parental help-seeking intentions |  |  |  |  | 0.75** | 0.60–0.92 |
| Child intelligence quotient |  |  |  |  | 1.01** | 1.00–1.02 |
| Parent-rated emotional symptoms |  |  |  |  | 0.98 | 0.92–1.03 |

\* $p < 0.05$ , \*\* $p < 0.01$ , \*\*\* $p < 0.001$ .
Abbreviations: CI, confidence intervals; OR, odds ratio; AQ, Autism-Spectrum Quotient; ref, reference.

## Discussion

Using a population-based cohort, this study examined the frequency of help-seeking intentions and preferences according to the level of autistic traits in adolescents and explored the mediating role of help-seeking intentions on adolescent depressive symptoms. In early adolescence (i.e., age 12), 40% of the adolescents with high autistic traits in the general population reported not having help-seeking intentions. This was despite the fact that 93% of them recognised the need for help and 97% of them had someone to ask for help. The respective percentages for those in the AQlow group were 27 %, 95%, and 98%. Although significantly fewer adolescents in the AQhigh group reported friends as their help-seeking source, the main source of help-seeking for both groups were their family. Our multivariable analysis confirmed that adolescents with high autistic traits were 1.84 times more likely to not to have help-seeking intentions compared with those with lower autistic traits. Help-seeking intentions at age 12 explained approximately 19% of the association between high autistic traits and depressive symptoms at age 14.

To our knowledge, this is the first study to systematically assess the association between autistic traits, help-seeking intentions, and subsequent depressive symptoms. Our results support our hypothesis that adolescents with high autistic traits would show less help-seeking intentions which, in turn, partially explained their increased risk for subsequent depressive symptoms. Our findings were consistent with self-reports of difficulty or reluctancy towards seeking help for mental health problems among adults with autism. (7, 8) We confirmed that, in the community setting, this lack of help-seeking intention was already present in early adolescence (i.e., by age 12) among adolescents with high autistic traits.

The lack of help-seeking intentions among adolescents with high autistic traits was not a result of their inability to recognise the problem, as most of them were able to identify the need for mental health support in the vignette case. It was also not due to the lack of a source on which they could rely for help, as adolescents from both groups reported similar, adequate accessibility to someone they could rely on. Previous qualitative studies reported that autistic adults had difficulties expressing their emotional distress to others, negative views on help-seeking based on their past experiences, and concerns over stigma attached to mental illness. (7, 8) Investigating into these possible mechanisms was beyond the scope of our study; however, these factors could have played a role in forming attitudes toward help-seeking among adolescents with high autistic traits. Clarifying the pathway to decreased help-seeking intentions among adolescents with high autistic traits will offer additional implications as to when and how tailored interventions can be implemented to promote help-seeking in this population.

As reported in the general population, (20) we found that the main source of help that was sought by adolescents with high autistic traits was their family. Previous studies have shown the importance of families as help-seeking partners throughout childhood and adolescence. (20, 21) For adolescents with high autistic traits, the role of the family as help-seeking partners may be more important than those in the lower range, as they reported significantly less help-seeking towards their friends, another main source of help-seeking partner during adolescence.

Importantly, in our study, parents of adolescents with high autistic traits also showed significantly less help-seeking intentions compared to parents of those in the lower range group. This may correspond with the results of a recent systematic review that reported more avoidance and less social support-seeking as coping strategies among parents of children with autism compared to parents of typically developing children. (22) Given that parental help-seeking intentions had a significant effect on the adolescents’ help-seeking intentions and that parents are often the gatekeepers to formal support for their children, (23) offering parents of adolescents with high autistic traits information regarding when and where to ask for mental health support and empowering them to seek help may be beneficial for the child.

In our study, help-seeking intentions mediated approximately 19% of the association between high autistic traits and depressive symptoms at age 14. Despite many studies focusing on help-seeking as a target for preventive interventions, the evidence from previous studies on help-seeking and mental health problems were mostly based on cross-sectional studies (9,10) and offered limited evidence for the longitudinal effects of help-seeking intentions on subsequent mental health problems. Although the effect is modest, our results suggest that promoting help-seeking intentions among children with high autistic traits may help reduce their risk of developing depressive symptoms during adolescence. This is inferred from a recent pilot randomised control trial of a school-based mental health literacy program that reported the effectiveness of their modified programs in meeting the needs of students with developmental disorders, including autism. (24) Given that individuals with high autistic traits already express fewer help-seeking intentions in early adolescence, ideally, the relevant interventions should be provided from before the children enter adolescence by modifying the interventions to ensure age-appropriate contents.

### Strengths and limitations

Our study had many strengths, which included the use of a population-based cohort. This allowed us to compare the results according to the level of autistic traits, adopt a prospective longitudinal design, and use self-reports from adolescents to assess help-seeking intentions and depressive symptoms. In our study, the participant’s autistic traits were measured by their parent, which reduced the possibility of shared method variance.

However, our study also had several limitations. First, although there were no differences in the proportion of each sex in our models, this may have been due to the small number of girls with high autistic traits that were included in our study. However, the proportion of girls with high autistic traits in our study was similar to those found in other population-based studies. (5) Second, over 90% of the adolescents with high autistic traits had intellectual abilities that were within the typical range (results provided upon request), indicating that our results may not apply to adolescents with high autistic traits and cognitive delay. Nevertheless, autistic individuals that have no cognitive delay are more prone to mental health problems (25) and are more likely to remain undiagnosed than their counterparts. (26) Thus, we propose that our community-based findings are important despite this limitation. Third, adolescents with high autistic traits may have experienced difficulty when interpreting the vignette or articulating their depressive symptoms. (27) However, almost all the adolescents from both groups recognised the presence of mental health problems and the need for help in the vignette. Moreover, the SMFQ has reliably been used among autistic adolescents in previous studies. (5) Finally, we did not have information on the actual help-seeking behaviours (i.e., actual service use) of the adolescents at age 14, which may have influenced the adolescents’ depressive symptoms at age 14. However, this is likely to lie on our hypothesised pathway between help-seeking intentions and subsequent depressive symptoms, as the theory of planned behaviour asserts that help-seeking intentions are a determinant of behaviour and this has been shown to correlate with actual service use. (28)

### Clinical Implications

Healthcare and educational practitioners should be aware that by the time they are on the cusp of adolescence, individuals with high autistic traits are already at increased risk of not having help-seeking intentions for their mental health problems, despite being at greater risk for mental health difficulties than their counterparts. In addition, parents of adolescents with high autistic traits also showed decreased help-seeking intentions. Thus, they require both attention and support as they are often the gatekeeper for the child’s access to formal mental health support. Promoting help-seeking intentions among these vulnerable adolescents through interventions adapted to meet their needs may help reduce their subsequent mental health problems. Further studies that clarify the underlying mechanism between high autistic traits, help-seeking intentions, and subsequent mental health problems will provide additional evidence for the development of these tailored interventions.

## Data Availability

The data that support the findings of this study are available on request from the corresponding author (MH). The data are not publicly available due to their containing information that could compromise the privacy of research participants.

## Declaration of Interest

None

## Funding

This study was supported by JSPS KAKENHI (KK, Grant Number JP16H06395, JP16H06399, JP16K21720), (SA, JP19H04877). However, none of the funders bear any responsibility for the analysis or interpretation of these data.

## Acknowledgements

We thank the Tokyo Teen Cohort (TTC) families for their time and cooperation, as well as the TTC study team for the use of data.

## Author Contribution

MH, SY, AN conceived the study. MH conducted the analysis and drafted the initial manuscript. All the authors revised and reviewed the manuscript. All authors approved the final manuscript as submitted.

## Ethical approval

All procedures involving human participants were approved by the institutional review boards of the Tokyo Metropolitan Institute of Medical Science (approval number: 12–35), SOKENDAI (Graduate University for Advanced Studies, 2012002), and the University of Tokyo (10057). Written informed consent was obtained from all the parents of the participating children, and informed assent was obtained from the children.

## Abbreviations

AQ: Autism-Spectrum Quotient
TTC: Tokyo Teen Cohort
SMFQ: Short Mood and Feelings Questionnaire
OR: odds ratio
CI: confidence interval

## Notes

### Competing Interest Statement

The authors have declared no competing interest.

### Author Declarations

All procedures involving human participants were approved by the institutional review boards of the Tokyo Metropolitan Institute of Medical Science (approval number: 12-35), SOKENDAI (Graduate University for Advanced Studies, 2012002), and the University of Tokyo (10057). Written informed consent was obtained from all the parents of the participating children, and informed assent was obtained from the children.

## Reference

1. American Psychiatric Association. Diagnostic and Statistical Manual of Mental Disorders, Fifth Edition. Association AP, editor. Washington,DC: APA Press; 2013.

2. Robinson EB, Koenen KC, McCormick MC, Munir K, Hallett V, Happe F, et al. Evidence that autistic traits show the same etiology in the general population and at the quantitative extremes (5%, 2.5%, and 1%). Arch Gen Psychiatry. 2011;68(11):1113–21.

3. Simonoff E, Pickles A, Charman T, Chandler S, Loucas T, Baird G. Psychiatric disorders in children with autism spectrum disorders: prevalence, comorbidity, and associated factors in a population-derived sample. J Am Acad Child Adolesc Psychiatry. 2008;47(8):921–9.

4. Gotham K, Brunwasser SM, Lord C. Depressive and anxiety symptom trajectories from school age through young adulthood in samples with autism spectrum disorder and developmental delay. J Am Acad Child Adolesc Psychiatry. 2015;54(5):369–76 e3.

5. Rai D, Culpin I, Heuvelman H, Magnusson CMK, Carpenter P, Jones HJ, et al. Association of Autistic Traits With Depression From Childhood to Age 18 Years. JAMA Psychiatry. 2018;75(8):835–43.

6. Patel V, Flisher AJ, Hetrick S, McGorry P. Mental health of young people: a global public-health challenge. The Lancet. 2007;369(9569):1302–13.

7. Crane L, Adams F, Harper G, Welch J, Pellicano E. ‘Something needs to change’: Mental health experiences of young autistic adults in England. Autism. 2019;23(2):477–93.

8. Coleman-Fountain E, Buckley C, Beresford B. Improving mental health in autistic young adults: a qualitative study exploring help-seeking barriers in UK primary care. Br J Gen Pract. 2020;70(694):e356–e63.

9. Aguirre Velasco A, Cruz ISS, Billings J, Jimenez M, Rowe S. What are the barriers, facilitators and interventions targeting help-seeking behaviours for common mental health problems in adolescents? A systematic review. BMC Psychiatry. 2020;20(1):293.

10. Singh S, Zaki RA, Farid NDN. A systematic review of depression literacy: Knowledge, help-seeking and stigmatising attitudes among adolescents. J Adolesc. 2019;74:154–72.

11. Ando S, Nishida A, Yamasaki S, Koike S, Morimoto Y, Hoshino A, et al. Cohort Profile: The Tokyo Teen Cohort study (TTC). Int J Epidemiol. 2019.

12. Jorm AF, Wright A, Morgan AJ. Beliefs about appropriate first aid for young people with mental disorders: findings from an Australian national survey of youth and parents. Early Interv Psychiatry. 2007;1(1):61–70.

13. Ando S, Nishida A, Usami S, Koike S, Yamasaki S, Kanata S, et al. Help-seeking intention for depression in early adolescents: Associated factors and sex differences. J Affect Disord. 2018;238:359–65.

14. Allison C, Auyeung B, Baron-Cohen S. Toward brief “Red Flags” for autism screening: The Short Autism Spectrum Quotient and the Short Quantitative Checklist for Autism in toddlers in 1,000 cases and 3,000 controls [corrected]. J Am Acad Child Adolesc Psychiatry. 2012;51(2):202–12 e7.

15. Angold A, Costello E, Messer S. Development of a short questionnaire for use in epidemiological studies of depression in children and adolescents. International Journal of Methods in Psychiatric Research. 1995;5:237–49.

16. Thapar A, McGuffin P. Validity of the shortened Mood and Feelings Questionnaire in a community sample of children and adolescents: a preliminary research note. Psychiatry Res. 1998;81(2):259–68.

17. Yamasaki S, Ando S, Richards M, Hatch SL, Koike S, Fujikawa S, et al. Maternal diabetes in early pregnancy, and psychotic experiences and depressive symptoms in 10-year-old offspring: A population-based birth cohort study. Schizophr Res. 2019;206:52–7.

18. Goodman R. The Strengths and Difficulties Questionnaire: a research note. J Child Psychol Psychiatry. 1997;38(5):581–6.

19. White IR, Royston P, Wood AM. Multiple imputation using chained equations: Issues and guidance for practice. Stat Med. 2011;30(4):377–99.

20. Rickwood DJ, Mazzer KR, Telford NR. Social influences on seeking help from mental health services, in-person and online, during adolescence and young adulthood. BMC Psychiatry. 2015;15:40.

21. Zwaanswijk M, van der Ende J, Verhaak PF, Bensing JM, Verhulst FC. Help-seeking for child psychopathology: pathways to informal and professional services in the Netherlands. J Am Acad Child Adolesc Psychiatry. 2005;44(12):1292–300.

22. Vernhet C, Dellapiazza F, Blanc N, Cousson-Gelie F, Miot S, Roeyers H, et al. Coping strategies of parents of children with autism spectrum disorder: a systematic review. Eur Child Adolesc Psychiatry. 2019;28(6):747–58.

23. Sayal K, Tischler V, Coope C, Robotham S, Ashworth M, Day C, et al. Parental help-seeking in primary care for child and adolescent mental health concerns: qualitative study. Br J Psychiatry. 2010;197(6):476–81.

24. Katz J, Knight V, Mercer SH, Skinner SY. Effects of a Universal School-Based Mental Health Program on the Self-concept, Coping Skills, and Perceptions of Social Support of Students with Developmental Disabilities. J Autism Dev Disord. 2020;50(11):4069–84.

25. Rai D, Heuvelman H, Dalman C, Culpin I, Lundberg M, Carpenter P, et al. Association Between Autism Spectrum Disorders With or Without Intellectual Disability and Depression in Young Adulthood. JAMA Netw Open. 2018;1(4):e181465.

26. Hosozawa M, Sacker A, Mandy W, Midouhas E, Flouri E, Cable N. Determinants of an autism spectrum disorder diagnosis in childhood and adolescence: Evidence from the UK Millennium Cohort Study. Autism. 2020;24(6):1557–65.

27. Hill E, Berthoz S, Frith U. Brief report: cognitive processing of own emotions in individuals with autistic spectrum disorder and in their relatives. J Autism Dev Disord. 2004;34(2):229–35.

28. Li W, Denson LA, Dorstyn DS. Understanding Australian university students’ mental health help-seeking: An empirical and theoretical investigation. Australian Journal of Psychology. 2018;70(1):30–40.

