## Supplementary material for "Lower help-seeking intentions and subsequent depressive symptoms among adolescents with high autistic traits: A population-based cohort study": Table S

Table S1 Descriptive characteristics according to the level of autistic traits *in the observed sample* (N = 2,505)

| | AQ low[a] (n=2,305, 92.0%) | | AQ high[a] (n=200, 8.0%) | | |
|---|---|---|---|---|---|
| | n | % | n | % | *p*-value[b] |
| Autistic traits at age 12, mean (SD) | 2,305 | 2.1 (1.3) | 200 | 6.7 (0.9) | < 0.001 |
| Sex | | | | | |
| Male | 1,189 | 51.6 | 136 | 68.0 | < 0.001 |
| Female | 1,116 | 48.4 | 64 | 32.0 | |
| Low annual household income[c] | | | | | |
| No | 1,997 | 89.6 | 167 | 88.4 | 0.60 |
| Yes | 232 | 10.4 | 22 | 11.6 | |
| Low maternal education[d] | | | | | |
| No | 1,924 | 84.0 | 161 | 81.3 | 0.33 |
| Yes | 367 | 16.0 | 37 | 18.7 | |
| Parental help-seeking intentions[e] | | | | | |
| No | 470 | 20.5 | 58 | 29.0 | 0.005 |
| Yes | 1,826 | 79.5 | 142 | 71.0 | |
| Child intelligence quotient, mean (SD) | 2,305 | 108.1 (13.9) | 199 | 106.8 (15.1) | 0.23 |
| Parent-rated emotional symptoms, mean (SD) | 2,297 | 1.5 (1.6) | 200 | 2.5 (2.3) | < 0.001 |
| Parent-reported diagnosis of autism by age 12 | | | | | |
| No | 2,276 | 99.4 | 183 | 92.4 | < 0.001 |
| Yes | 15 | 0.7 | 15 | 7.6 | |
| Self-reported depressive symptoms at age 14, mean (SD) | 1,691 | 3.0 (4.6) | 144 | 4.1 (5.6) | 0.007 |

Variables were measured in the first wave of the study (age 10) unless otherwise stated.

[a] Defined as scoring above the suggested clinical-cut off on the short form of the Autism-Spectrum Quotient adolescent version measured by a parent at age 12.

[b] *p*-value for group difference obtained from Chi-square tests for categorical variables and from t-tests for continuous variables.

[c] Defined as a household income below 4,000,000 Yen.

[d] Defined as a high school graduate or below.

[e] Measured through the adult version of the depression vignette.

Abbreviations: AQ, Autism-Spectrum Quotient; SD, standard deviation.

Table S2 Sample bias analysis

| | Total TTC sample (N = 3,171) | | | | |
|---|---|---|---|---|---|
| | Analytic sample (n =2,505) | | Non-analytic sample[a] (n = 666) | | |
| | n | % | n | % | $p$-value[b] |
| Autistic traits, mean (SD) | 2,505 | 2.5 (1.8) | 237 | 2.7 (1.8) | 0.21 |
| Sex | | | | | |
| Male | 1,325 | 52.9 | 359 | 53.9 | 0.64 |
| Female | 1,180 | 47.1 | 307 | 46.1 | |
| Low annual household income | | | | | |
| No | 2,164 | 89.5 | 557 | 88.7 | 0.56 |
| Yes | 254 | 10.5 | 71 | 11.3 | |
| Low maternal education | | | | | |
| No | 2,085 | 83.8 | 537 | 81.7 | 0.21 |
| Yes | 404 | 16.2 | 120 | 18.3 | |
| Parental help-seeking intentions | | | | | |
| No | 528 | 21.2 | 148 | 22.4 | 0.49 |
| Yes | 1,968 | 78.9 | 513 | 77.6 | |
| Child intelligence quotient, mean (SD) | 2,504 | 108.0 (14.0) | 664 | 106.8 (14.6) | 0.05 |
| Parent-rated emotional symptoms, mean (SD) | 2,497 | 1.6 (1.5) | 663 | 1.7 (1.7) | 0.08 |
| Parent-reported diagnosis of autism by age 12 | | | | | |
| No | 2,459 | 98.8 | 480 | 97.4 | 0.02 |
| Yes | 30 | 1.2 | 13 | 2.6 | |
| Child not having help-seeking intention | | | | | |
| No | 1,814 | 72.4 | 16 | 72.7 | 0.97 |
| Yes | 691 | 27.6 | 6 | 27.3 | |
| Self-reported depressive symptoms at age 14, mean (SD) | 1,853 | 2.5 (4.0) | 191 | 3.1 (4.7) | 0.06 |

[a]Children excluded from this study due to non-participation at wave 2 (age12), missing data for autistic traits or child's help-seeking intention.

[b]$p$-value for group difference obtained from the Chi-square tests for categorical variables and from t-tests for continuous variables.

Abbreviations: SD, standard deviation.

Table S3 Depressive symptoms at age 14 by level of autistic traits

| Depressive symptoms | Model 1 Crude | Model 2 Sex adjusted | Model 3 Confounder adjusted[a] |
|---|---|---|---|
| | b (95% CI) | b (95% CI) | b (95% CI) |
| AQlow | ref | ref | ref |
| AQhigh | 1.02 (0.28–1.77)** | 1.29 (0.56–2.03)** | 1.04 (0.30–1.78)** |

*$p < 0\ .05$, **$p < 0.01$.[a] Further adjusted for low annual household income, low maternal education, parental help-seeking intention, the adolescent's intelligence quotient and parent-rated emotional symptoms all measured at age 10.

Abbreviations: AQ, Autism-Spectrum Quotient.

Table S4 Estimates of direct and indirect effect (mediated through adolescent's help-seeking intention) in the association between autistic traits and depressive symptoms at age 14

| | Indirect effect | Direct effect | Total effect |
|---|---|---|---|
| Depressive symptoms | b (95% CI) | b (95% CI) | b (95% CI) |
| AQ low | ref | ref | Ref |
| AQ high | 0.20 (0.06–0.33)** | 0.84 (0.18–1.51)* | 1.04 (0.37–1.71)** |

*$p < 0$ .05, **$p < 0.01$. Mediating effect examined using structural equation modelling in Mplus adjusted for sex, low annual household income, low maternal education, parental help-seeking intention, the adolescent's intelligence quotient and parent-rated emotional symptoms are shown.

Abbreviations: AQ, Autism-Spectrum Quotient.

Table S5 Adolescent's help-seeking intention and depressive symptoms by level of autistic traits using *observed cases*

| | Not having help-seeking intention (N = 2,387) | | Depressive symptoms (N = 1,753) | |
|---|---|---|---|---|
| | Model 3 Confounder adjusted[a] | | Model 3 Confounder adjusted[a] | |
| | OR | 95% CI | b | 95% CI |
| AQ high (ref = AQ low) | 1.84*** | 1.34–2.53 | 1.15** | 0.34–1.96 |
| Sex (ref = female) | 1.13 | 0.94–1.36 | -1.76*** | -2.19– -1.33 |
| Low annual household income | 1.03 | 0.76–1.39 | 0.68 | -0.05–1.41 |
| Low maternal education | 1.12 | 0.87–1.44 | 0.23 | -0.39–0.84 |
| Parental help-seeking intentions | 0.75** | 0.60–0.93 | -0.70** | -1.22– -0.18 |
| Child intelligence quotient | 1.01* | 1.00–1.02 | 0.00 | -0.01–0.02 |
| Parent-rated emotional symptoms | 0.98 | 0.93–1.03 | 0.21** | 0.08–0.34 |

*$p$ < 0 .05, **$p$ < 0.01, ***$p$ < 0.001. [a]Adjusted for sex, low annual household income, low maternal education, parental help-seeking intentions, the adolescent's intelligence quotient and parent-rated emotional symptoms all measured at age 10.

Abbreviations: AQ, Autism-Spectrum Quotient.

Table S6 Adolescent's help-seeking intentions and depressive symptoms by level of autistic traits, *without children diagnosed as autism* (N = 2,475)

| | Not having help-seeking intentions | | Depressive symptoms | |
|---|---|---|---|---|
| | Model 3<br>Confounder adjusted[a] | | Model 3<br>Confounder adjusted[a] | |
| | OR | 95% CI | b | 95% CI |
| AQ high (ref = AQ low) | 1.93*** | 1.41–2.64 | 0.80* | 0.08–1.52 |
| Sex (ref = female) | 1.13 | 0.94–1.35 | -1.64*** | -2.01– -1.26 |
| Low annual household income | 1.09 | 0.81–1.47 | 0.67* | 0.03–1.31 |
| Low maternal education | 1.12 | 0.88–1.44 | 0.29 | -0.24–0.83 |
| Parental help-seeking intentions | 0.76* | 0.62–0.94 | -0.54* | -1.02– -0.06 |
| Child intelligence quotient | 1.01** | 1.00–1.02 | 0.00 | -0.01–0.02 |
| Parent-rated emotional symptoms | 0.97 | 0.92–1.02 | 0.20** | 0.08–0.31 |

*$p$ < 0 .05, **$p$ < 0.01, ***$p$ < 0.001. [a]Adjusted for sex, low annual household income, low maternal education, parental help-seeking intentions, the adolescent's intelligence quotient and parent-rated emotional symptoms all measured at age 10.

Table S7 Estimates of direct and indirect effect (mediated through help-seeking intentions) in the association between autistic traits and depressive symptoms at age 14, *without children diagnosed as autism* (N = 2,475)

| | Indirect effect | Direct effect | Total effect |
|---|---|---|---|
| Depressive symptoms | b (95% CI) | b (95% CI) | b (95% CI) |
| AQ low | ref | ref | ref |
| AQhigh | 0.20 (0.06–0.33)** | 0.61 (-0.06–1.27) | 0.80 (0.14–1.46)* |

*$p < 0.05$, **$p < 0.01$. Mediating effect examined using structural equation modelling in Mplus adjusted for sex, low annual household income, low maternal education, parental help-seeking intentions, the adolescent's intelligence quotient and parent-rated emotional symptoms are shown.
